# Aberrant neurophysiological signaling underlies speech impairments in Parkinson’s disease

**DOI:** 10.1101/2022.04.01.22273315

**Authors:** Alex I. Wiesman, Peter W. Donhauser, Clotilde Degroot, Sabrina Diab, Shanna Kousaie, Edward A. Fon, Denise Klein, Sylvain Baillet, PREVENT-AD Research Group, Quebec Parkinson Network

## Abstract

Difficulty producing intelligible speech is a common and debilitating symptom of Parkinson’s disease (PD). Yet, both the robust evaluation of speech impairments and the identification of the affected brain systems are challenging. We examine the spectral and spatial definitions of the functional neuropathology underlying reduced speech quality in patients with PD using a new approach to characterize speech impairments and a novel brain-imaging marker. We found that the interactive scoring of speech impairments in PD (N=59) is reliable across non-expert raters, and better related to the hallmark motor and cognitive impairments of PD than automatically-extracted acoustical features. By relating these speech impairment ratings to neurophysiological deviations from healthy adults (N=65), we show that articulation impairments in patients with PD are robustly predicted from aberrant activity in the left inferior frontal cortex, and that functional connectivity of this region with somatomotor cortices mediates the influence of cognitive decline on speech deficits.

---

Parkinson’s disease (PD) is the second most common neurodegenerative disorder worldwide^1^, and is characterized by progressive declines in motor function and cognition. Difficulties producing intelligible speech are some of the earliest^2-4^ and most debilitating^5,6^ impairments of PD. Speech production is inherently complex, and speech symptoms in patients with PD are multidimensional^7^; they commonly include hoarse *voice*, imprecise *articulation*, and monotonous *prosody*^4,8^. However, the best approach to quantifying pathological changes in speech remains unclear. On the one hand, evaluation of dysarthric symptoms by a certified speech-language pathologist using clinical scales is time-consuming but often regarded as the gold standard. On the other, more rapid and inexpensive assessments based on automatically-extracted acoustical measures^4,9,10^ are promising, but may not provide a sufficiently nuanced representation of the disease symptomatology. The optimal outcome of remediation speech therapies in PD is improved intelligibility to human listeners, hence the value of human assessment of speech impairments as an alternative. Human ratings of speech impairments in PD are highly reliable across raters^11-13^, time points^14^, and levels of rater expertise^13^, and can contribute to predicting disease progression^12^ and therapeutic outcomes^14^ beyond simpler acoustical metrics.

Beyond the robust qualification of speech impairments in PD, their neurophysiological origins are also unknown. In the healthy brain, speech production engages a distributed and predominantly left-lateralized ensemble of cortical regions including the primary motor, primary auditory, pre-motor, posterior parietal and inferior frontal cortices^15-21^. Temporally, cued speech production requires the integration of incoming phonological information in the left superior temporal cortex, which precedes and then overlaps with sustained motor processing in the precentral gyrus, followed by engagement of left inferior frontal cortex (LIFC) for metrical encoding of speech production, and the left middle and superior frontal cortices for voluntary control of motor initiation^18,22-25^. The frequency-specific (spectral) components of neurophysiological activity related to speech production have also been studied extensively^26,27^. They include a key role for alpha (∼7–13 Hz) and beta (∼15–30 Hz) frequency bands in speech-network regions, and slower delta band (∼2-4 Hz) activity in prefrontal regions^28,29^, for effective production of speech^28,30-35^. Interregional beta-band functional connectivity between prefrontal, auditory, and motor cortices is also essential for healthy speech production^28,36^.

Functional neuroimaging studies of patients with PD have shown that speech production recruits greater cerebral blood flow and oxygenation across prefrontal, auditory, and motor regions^37-40^ than in healthy controls, and that this hypermetabolism is normalized by common PD therapies^37,38^. Inter-regional connectivity of the speech circuit also appears to impact speech production in PD^41,42^, with opposing effects of decreased versus increased functional connectivity during speech preparation and production, respectively^43^. The neurophysiological spectrum of these effects is less studied; a limited literature suggests decreased beta oscillations in the subthalamic nucleus^44^ and primary motor cortex^45^ in PD patients during active speech. Importantly, it remains relatively unclear to what extent these patterns of aberrant neural activity during active speech production under highly controlled experimental conditions relate to the real-world difficulties experienced by patients with PD. Given the key role of rhythmic neural activity in motor and cognitive impairments in patients with PD^46-48^, and the proven and future potential for therapeutic interventions based on frequency-specific neurostimulation in this population^49-51^, a clearer understanding of the spatio-spectral neural bases of speech intelligibility deficits in PD is essential.

In this study, we examine the spectral and spatial definitions of the functional neuropathology underlying reduced speech quality in patients with PD (N = 59). Towards this goal, we first quantified patient speech impairments with a novel interactive tool designed for non-specialists, and show that this approach outperforms measures based on the automatic extraction of acoustical features. We then introduce a new brain mapping technique of spectral neurophysiological deviations (the Spectral Deviation Index; SDI) between each patient and a group of demographically-matched healthy controls (N = 65). We hypothesized that aberrant neurophysiological manifestations would map to brain regions known for their involvement in speech production, and scale with the severity of speech impairments in patients with PD. We also anticipated that neurophysiological connectivity across the brain circuit for speech production would also be altered in those patients with more pronounced speech difficulties.

## Results

### Demographics, clinical assessments, and speech impairments in patients with PD

Demographics for both groups, as well as clinical features for the PD group, are reported in Table 1. Speech quality assessments were consistent across raters for all assessed features: voice (intra-class correlation coefficient [ICC] = .89, 95% CI = [.83 .93]), articulation (ICC = .88, 95% CI = [.81 .92]), and prosody (ICC = .78, 95% CI = [.66 .86]; Fig. 1A). These speech impairment ratings were also related to clinical features of PD (Fig. 1B): they were all significantly predicted by clinical motor function (UPDRS-III, minus speech sub-scores; voice: *t*(51) = 2.82, *p* = .007; articulation: *t*(51) = 3.61, *p* < .001; prosody: *t*(51) = 3.07, *p* = .003), such that greater non-speech motor impairment predicted greater speech difficulties. We found weaker associations between all three speech features and PD staging (Hoehn & Yahr scale; voice: *t*(45) = 2.05, *p* = .046; articulation: *t*(45) = 2.07, *p* = .044; prosody: *t*(45) = 2.05, *p* = .046; all uncorrected for multiple comparisons). Only articulation impairments were predicted by patient cognitive abilities (MoCA scores; voice: *t*(56) = -0.47, *p* = .639; articulation: *t*(51) = -2.85, *p* = .006; prosody: *t*(51) = -1.15, *p* = .250), such that greater cognitive impairment was associated with greater articulation difficulties. Further, models including these speech ratings predicted non-speech motor dysfunction (ΔAIC = -6.69) and cognitive function (ΔAIC = -9.03) better than acoustical features extracted automatically from the same recordings. All three features also significantly predicted speech impairment ratings made by a trained clinician (UPDRS-III speech sub-score; voice: *t*(51) = 5.17, *p* < .001; articulation: *t*(51) = 3.51, *p* < .001; prosody: *t*(51) = 4.49, *p* < .001; Fig. S1).

**Table 1.** Between-group demographic comparisons and patient group clinical profile.

|  | Age<br>(years) | Sex<br>(% female) | Handedness<br>(# left/ambi) | Education<br>(years) |  |  |
| --- | --- | --- | --- | --- | --- | --- |
| HC | 63.02 (8.13) | 65 | 3/3 | 15.85 |  |  |
| PD | 65.95 (8.14) | 71 | 4/3 | 15.14 |  |  |
| *p* | .067 | .434 | .863 | .615 |  |  |
|  | MoCA | UPDRS-III | UPDRS-III<br>Speech<br>(N = 54) | Hoehn &<br>Yahr<br>(N = 48) | Years Since<br>Dx<br>(N = 56) | % Taking<br>DA Agonists<br>(N = 53) |
| PD |  |  |  |  |  |  |
| Range | 12 – 30 | 7 – 71 | 0 – 3 | 1 – 3 | 1 – 25 | - |
| Mean<br>(SD) | 24.44 (3.99) | 32.68 (14.77) | 1.35 (0.83) | 1.96 (0.70) | 6.66 (4.74) | 28.81 |

HC: healthy control group; PD: Parkinson's disease group; MoCA: Montreal Cognitive Assessment; UPDRS-III: Unified Parkinson's Disease Rating Scale part III; Dx: Diagnosis; DA: Dopamine. Unless otherwise indicated, values indicate means and associated parentheticals indicate standard deviations. HC sample N = 65, PD sample N = 59, unless otherwise indicated. P-values indicate significance of between-groups Mann-Whitney U tests and chi-square tests for continuous and categorical variables, respectively.

**Figure 1.**
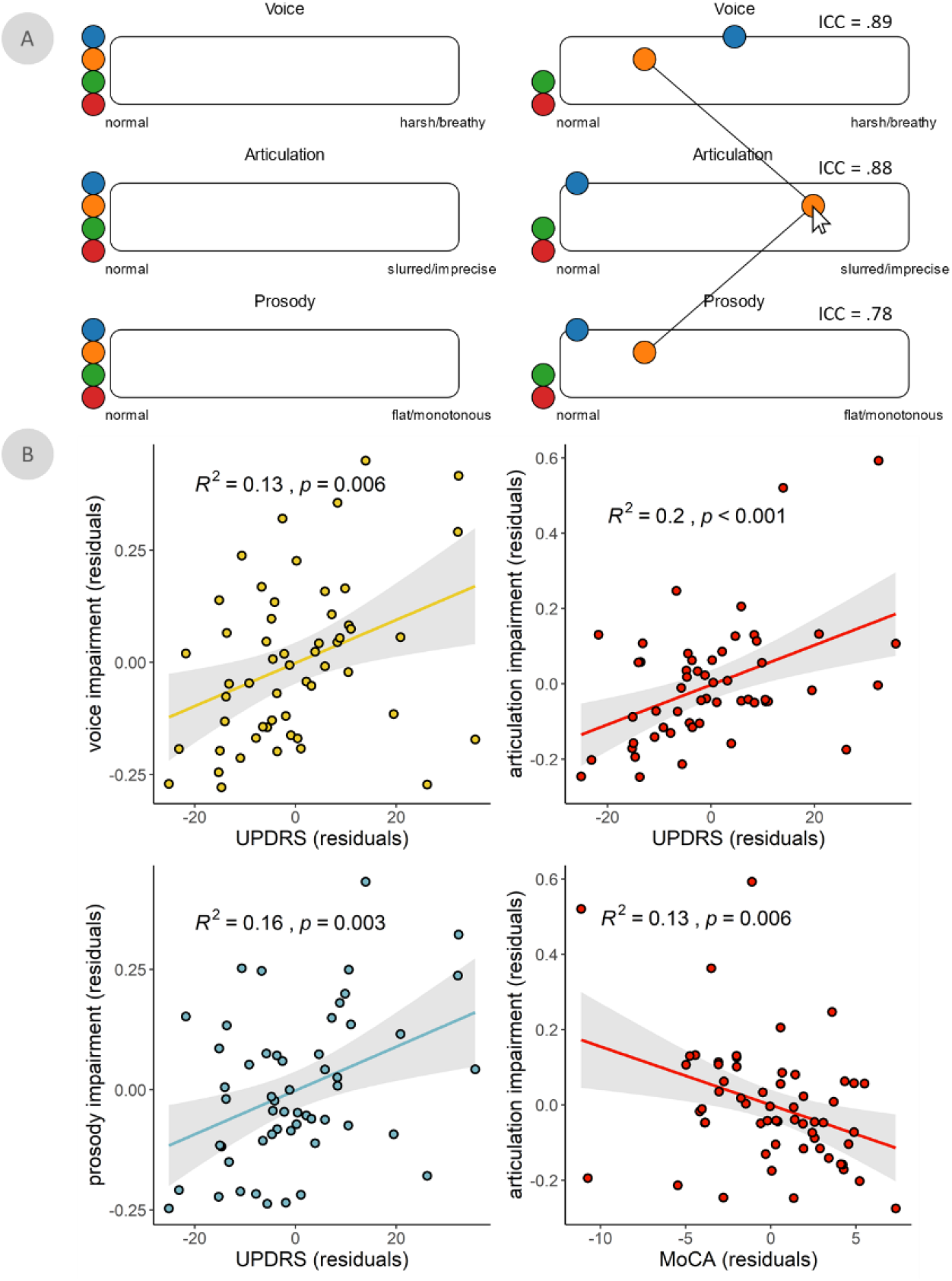
Speech impairment ratings are reliable and predicted by clinical features of PD. (A) Graphical user interface for the manual rating of speech feature impairments using *Audio-Tokens*. When hovered-over with the mouse, colored circles play speech samples from separate patients with Parkinson’s disease, allowing for interactive and comparative rating (i.e., by clicking and sliding the circles horizontally) of impairments in each speech feature. Intraclass correlation coefficients (ICC) indicate reliability of speech ratings across three independent raters. (B) Significant linear relationships between voice, articulation, and prosody impairments and two common clinical scales in Parkinson’s disease: the Unified Parkinson’s Disease Rating Scale part III (i.e., UPDRS-III, minus speech sub-scores) and the Montreal Cognitive Assessment (MoCA). All models controlled for age. Shaded intervals represent the 95% confidence interval.

### Spectral pathology is related to articulation impairments in PD

The average of individual SDI maps (Fig. 2A) across all patients emphasized spectral deviations in premotor, primary somatomotor, and superior parietal cortices bilaterally (Fig. 2B). The greatest variability of SDI across patients was found in the bilateral prefrontal and temporal cortices (Fig. 2C).

**Figure 2.**
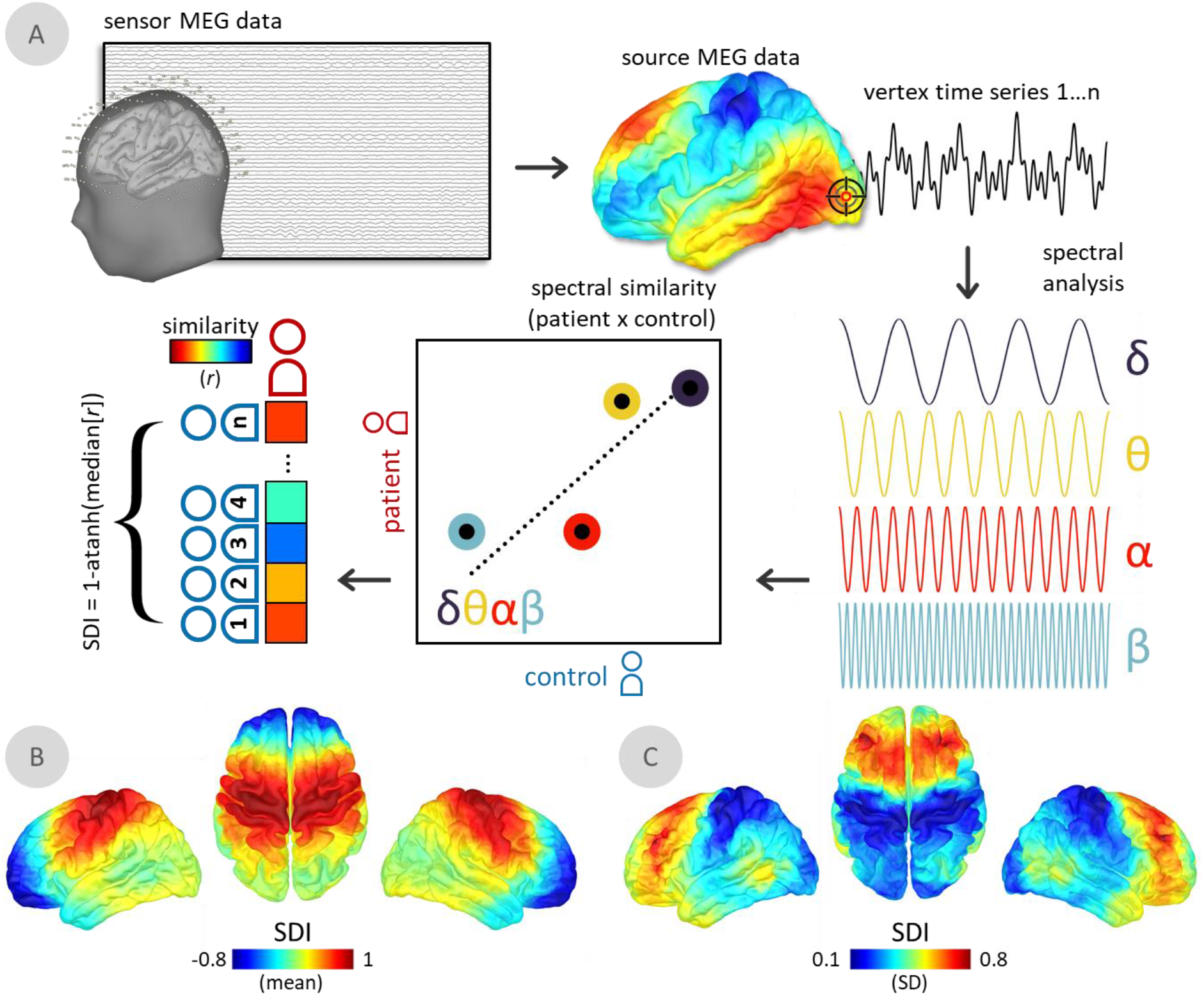
The Spectral Deviation Index (SDI). (A) For all participants, magnetoencephalography (MEG) data were cortically mapped, frequency-transformed, and averaged over canonical frequency bands (i.e., δ: 2 – 4 Hz; θ: 4 – 7 Hz; d: 8 – 12 Hz; β: 15 – 29 Hz). Pearson correlation coefficients were then estimated at each vertex, representing the similarity in frequency-wise power between each patient and control. For each patient, the median of these values was taken over all comparisons to control participants, which was then Fisher-transformed to ensure linear scaling (i.e., transformed with the inverse hyperbolic tangent function) and subtracted from 1 to indicate relative deviations from the control group. Surface maps below indicate the mean (B) and standard deviation (C) of the spectral deviation maps across all patients with Parkinson’s disease.

We performed a multiple regression of these maps on all three speech features, and found that articulation impairments uniquely predicted spectral pathology in the LIFC (TFCE; *p*FWE = .027; peak vertex = x: -51, y: 36, z: 2; Fig. 3A), with greater LIFC deviations associated with greater speech impairment (Fig. 3B). Neural activity in all tested frequency bands (from delta to beta) contributed to this effect (ΔAIC; delta = 10.56; theta = 2.86; alpha = 9.74; beta = 3.80; Fig. 3C), with the strongest influences from the delta and alpha bands. Post-hoc analysis of frequency-specific relationships suggested that preserved speech function was related to slowing of neural activity in the LIFC: greater impairment was associated with increased activity in the faster (alpha & beta) and decreased activity in the slower (delta & theta) frequency bands (Fig. 3D). SDI values in the LIFC did not significantly relate to clinical motor function (UPDRS-III, minus speech sub-scores; *t*(51) = 1.54, *p* = .130) or MoCA (*t*(56) = -0.25, *p* = .807) scores. The relationship between cross-spectral pathology and articulation impairment remained significant (*p* = .001) when potential confounds were added to the model, including head motion (*p* = .403), eye movements (*p* = .242), cardiac artifacts (*p* = .770), local cortical thickness (*p* = .717), and local aperiodic slope (*p* = .666).

**Figure 3.**
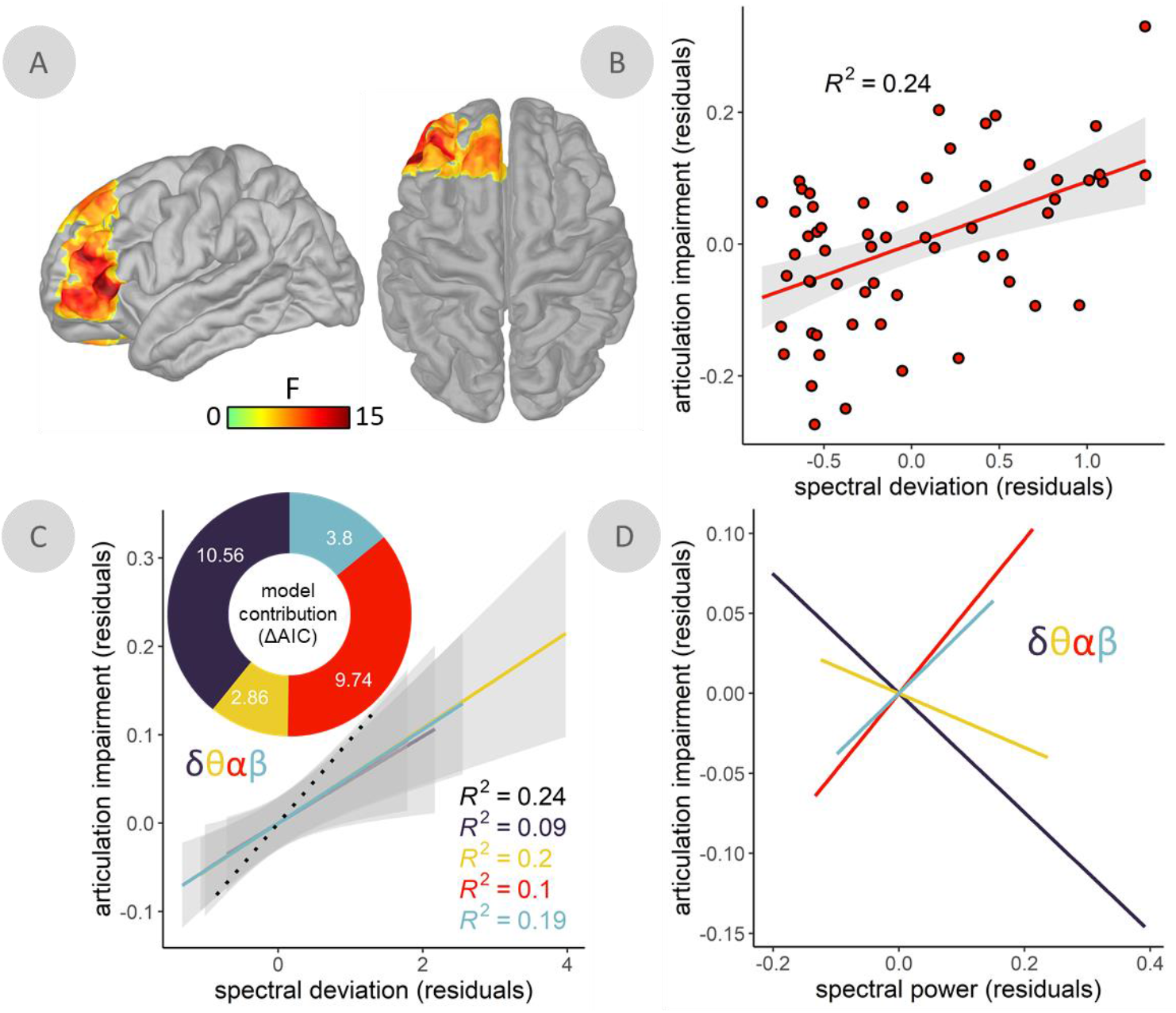
Spectral deviation predicts articulatory impairments in the left inferior frontal cortex. (A) Surface maps indicate a significant relationship between spectral deviations and articulation impairments in patients with Parkinson’s disease (PD), with spectral deviations (SDI) and articulation impairment ratings from the peak vertex of this relationship plotted in (B). (C) Model contribution values shown in the donut plot indicate comparisons between models predicting articulation impairment ratings using the SDI values from the same peak vertex and comparable SDI values computed with each frequency left out once. More positive differences in Akaike information criterion (ΔAIC) indicate a stronger contribution of that frequency to the overall model, with ΔAIC > 2 signifying meaningful differences in model information. Best fit lines represent the linear relationships between these leave-one-out models for each frequency and the articulation impairment ratings across patients with PD. (D) Lines-of-best-fit indicate the direction of the underlying relationships between spectral power at each frequency and articulation impairment ratings. All models controlled for age. Shaded intervals represent the 95% confidence interval.

### LIFC-somatomotor beta-frequency connectivity mediates cognitive contributions to articulation impairments in PD

We computed whole-cortex frequency-specific connectivity maps to examine whether signal similarities between the LIFC and the rest of the cortex were related to speech impairments in PD. The connectivity analyses were seeded at the vertex location corresponding to the peak of the SDI-articulation impairment statistical map (x: -51, y: 36, z: 2). We found that articulation impairment predicted connectivity between LIFC and a distributed network of pre-motor, anterior cingulate, and somatomotor regions in the beta band, above and beyond the effects of age, prosody impairments, and voice impairments (TFCE; *p*FWE < .001; peak vertex = x: -11, y: -12, z: 46; Fig. 4A). This relationship was such that patients with weaker LIFC-somatomotor functional connectivity exhibited worse articulation impairments (Fig. 4B, left). Further, this shared variance was independent of SDI effects in the LIFC, as both LIFC SDI values (*t*(56) = 4.28, *p* < .001) and beta LIFC-somatomotor connectivity (*t*(56) = -5.66, *p* < .001) each significantly predicted articulation impairment, above and beyond the other, when included in a single linear model. Beta LIFC-somatomotor functional connectivity did not relate to UPDRS III scores, but did significantly covary with MoCA scores (*t*(56) = 2.98, *p* = .004; Fig. 4B, right), such that stronger connectivity predicted better cognitive function. The relationship between MoCA scores and articulation impairment was also fully mediated by beta-band connectivity (causal mediation analysis, 10,000 simulations; Fig. 4C), as indicated by a significant indirect effect (average causal mediation effect = -.005, *p* = .024) and a non-significant direct effect (average direct effect = -.011, *p* = .223) upon addition of connectivity values to the linear model. The relationship between beta LIFC-somatomotor connectivity and articulation impairment remained significant (*p* < .001) when potential confounds were added to the model, including head motion (*p* = .750), eye movement (*p* = .451), and cardiac artifacts (*p* = .771).

**Figure 4.**
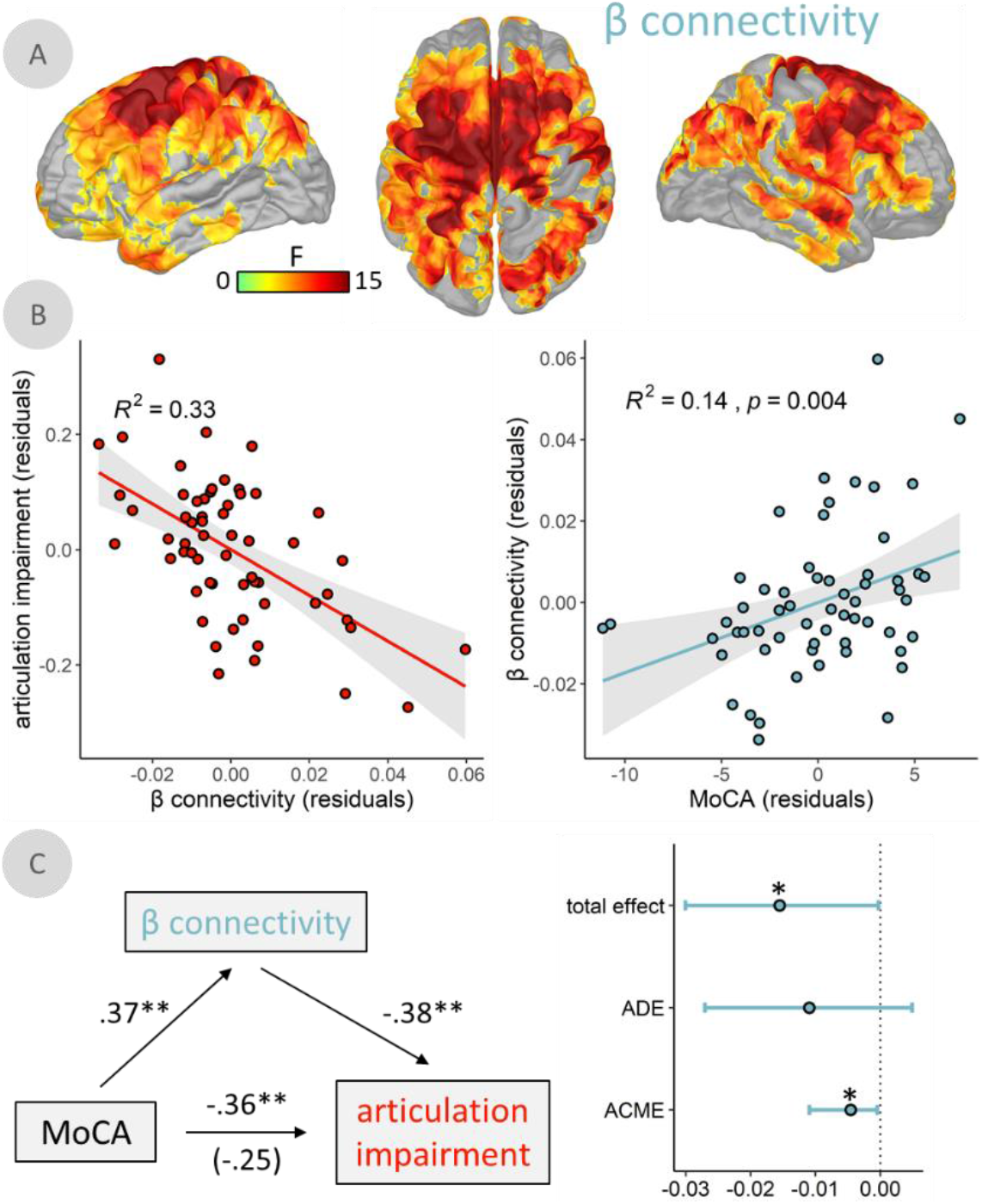
β-frequency functional connectivity between left inferior frontal cortex and somatomotor regions mediates the impact of cognitive decline on articulation impairments. (A) Surface maps indicate a significant relationship between articulation impairment ratings and frequency-resolved functional connectivity, computed using orthogonalized amplitude envelope correlations (AEC) with the peak vertex of the relationship from Figure 3 as the seed. (B) β-connectivity and articulation impairments from the peak vertex of this relationship are plotted on the left, and the relationship between the same β-connectivity values and cognitive function (i.e., Montreal Cognitive Assessment [MoCA] scores) plotted on the right. (C) Paths between MoCA scores, articulation impairment ratings, and β-connectivity values to the left indicate partial correlations (*r*) between each set of variables. The correlation value in parentheses represents the mediated relationship between MoCA scores and articulation impairment ratings when accounting for β-connectivity. The total effect, average direct effect (ADE), and average causal mediation effect (ACME; i.e., indirect effect) are plotted with 95% confidence intervals to the right. All models controlled for age. Shaded intervals represent the 95% confidence interval. \*\**p* < .01, \**p* < .05.

## Discussion

This study introduces and combines novel approaches to spectral brain mapping and speech impairment quantification to delineate the functional neural pathology contributing to speech impairment in PD. We identify in a large group of patients with PD a pathological relationship between articulation impairments and spectral deviations in the LIFC, with strongest contributions from neurophysiological activity in the delta and alpha bands. In healthy adults, the LIFC is a hub that exhibits multi-frequency interactions with a number of language network regions^29^. Our data also showed that neurophysiological connectivity between LIFC and a network of somatomotor cortices in the beta band independently predicted articulation impairments, and fully mediated the effect of cognitive abilities on these impairments. Together, these results provide a spatially- and spectrally-resolved cortical network underlying articulatory impairments in PD. These findings may be of significance to future biomarker research and therapeutic targeting in PD. We also anticipate that the new, individualized modeling approach of spectral brain pathology for each patient may translate and be meaningful to other clinical populations.

Our approach to speech impairment quantification is predicated on the notion that clinical endpoints for speech production studies in PD need to improve speech perception by human raters^11-13,52,53^. One novel contribution of the present study is rating of speech features from non-experts through an intuitive and interactive app (*Audio-Tokens*)^54^, that facilitates simultaneous comparison of speech samples from multiple study participants. Despite being obtained from non-experts, the ratings exhibited a degree of inter-rater reliability comparable to those from gold-standard procedures in the field, such as speech intelligibility quantification using transcription^53^. The *Audio-Tokens* ratings predicted both non-speech motor features (i.e., UPDRS-III with speech sub-scores subtracted) and cognitive function (i.e., MoCA scores) more effectively than common acoustical features derived from automated methods. The *Audio-Tokens* ratings obtained remotely from non-experts were also similar to the assessments performed in the clinic by a trained professional (i.e., the speech sub-score of the UPDRS-III). We believe these findings establish the potential utility of the *Audio-Tokens* approach in patients with PD, particularly since comparable speech samples can be easily collected and rated without in-person visits to the clinic. The significance is that of tele-medical, longitudinal patient evaluations. A thorough comparison to more advanced automated speech quality quantification procedures (e.g., based on machine-learning techniques)^55^ remains warranted, albeit beyond the scope of this study. If these comparisons favor our approach, the additional predictive clinical power of human speech ratings can be harnessed to improve automated speech extraction methods, which in turn can alleviate the costs of and limited access to movement disorder specialists.

By relating speech impairments to spectral deviations from healthy neurophysiological activity, we identified that the greater the spectral deviations in LIFC, the more pronounced the articulation deficits in patients with PD. The LIFC is a key node of the speech production network^15-17,56^ associated to the metrical encoding of to-be-produced speech representations^56,57^. Previous research^58,59^ also suggests that the anterior location of this effect along the inferior frontal gyrus might indicate a deficit in the semantic and/or lexical aspects of articulatory functions in PD. Further, the slower range of neurophysiological activity involved (delta & theta frequency bands) is associated to temporal expectation and parsing mechanisms of sensory inputs^27,60-63^. We interpret these observations as PD patients experiencing difficulties producing clear and precise speech sounds because of impaired mechasnisms for parsing and rhythmically encoding phonological information prior to speech motor initiation. We also found that increased expressions of faster frequencies (alpha & beta frequency bands) in LIFC predicted speech deficits. Strong alpha/beta activity in the parieto-occipital^64,65^ and somatomotor^66-68^ cortices reflects enhanced functional local inhibition. Additionally, alpha/beta activity in LIFC is reduced around speech preparation^34,69,70^. Taken together, we interpret our findings as indicative that patients with reduced inhibition of LIFC at rest exhibited the best articulation abilities, possibly because of greater ability to parse speech information in time. Overall, these effects involve at once several components of the neurophysiological frequency spectrum in a single brain region, highlighting the utility of the proposed spectral deviation approach.

In contrast to the multi-spectral and spatially-focal nature of speech-related neural pathology in the LIFC, the network-level connectivity patterns that predicted articulation abilities in patients with PD were limited to the beta-band and spatially-widespread. Our data show that beta-band functional connectivity between the LIFC and prefrontal, frontal, and parietal somatomotor regions was negatively associated with speech impairments in patients. Substantial structural and functional connectivity effects have been reported between these regions^71,72^, and functional connectivity between LIFC and superior frontal regions during speech comprehension is reduced in PD^73,74^. Previous studies have also reported increased beta connectivity in patients with PD^47^, with indications of clinical significance, although it is still unclear if the relationship is one of pathology^75,76^ or compensation^77-80^. Our results point at a possible compensation role for beta-band connectivity in speech production: in the tested patient cohort, the stronger the LIFC-somatomotor beta connectivity, the higher the cognitive and articulation abilities. This effect was statistically independent of the spectral deviations we observed in LIFC, which indicates that both effects represent distinct PD functional pathologies of the speech production brain systems. We hope that these findings can inspire future research of individualized clinical monitoring and interventions via, for example, non-invasive therapeutic neuromodulation. We also found that the strength of connectivity effects mediated the relationship between cognitive abilities (MoCA scores) and articulation impairments. However, we did not observe such an effect between motor impairments and speech deficits. Our interpretation is that reduced beta-band connectivity between LIFC and somatomotor cortices is a biological proxy for cognitive contributions to articulation deficits in PD.

In sum, we believe our data advance the understanding of basic mechanisms involved in speech production in health and disease. They also highlight two new dissociable neurophysiological markers of symptom-specific clinical decline in PD, which improve the targeting of non-invasive neuromodulatory therapies in PD^49,51^. Further, the principle of the SDI is generalizable to other neurological and/or psychiatric disorders. In particular, the combination of easily administered speech sample recordings with a short resting state MEG has potential for identifying biomarkers in a host of neurodegenerative and neurodevelopmental disorders, many of which have speech impairment as a presenting complaint.

## Methods

### Participants

The Research Ethics Board at the Montreal Neurological Institute reviewed and approved this study. Written informed consent was obtained from every participant following detailed description of the study, and all research protocols complied with the Declaration of Helsinki. Exclusionary criteria for all participants included current neurological (other than PD) or psychiatric disorder; MEG contraindications; and unusable MEG, speech sample, or demographic data. All participants completed the same speech and MEG protocols with the same instruments at the same site.

Patients with mild to moderate idiopathic PD were enrolled as a part of the Quebec Parkinson Network (QPN; https://rpq-qpn.ca/)^81^ initiative, which includes extensive clinical, neuroimaging, neuropsychological, and biological profiling for each participant. A final sample of 59 participants with PD fulfilled the criteria of having complete and useable MEG, speech sample, and demographic data. All patients with PD were prescribed a stable dosage of antiparkinsonian medication with satisfactory clinical response prior to study enrollment. Patients were instructed to take their medication as prescribed before research visits, and thus all data were collected in the practically-defined “ON” state. The motor subtest of the Unified Parkinson’s Disease Rating Scale (UPDRS-III)^82^ and Montreal Cognitive Assessment (MoCA)^83^ were administered to all participants. We also computed all relationships between motor pathology and speech feature ratings with the clinical speech sub-score (UPDRS-III, item 1; sub-scores available for N = 54) subtracted from the UPDRS-III scores to avoid bias. Additional clinical data were also available for subsamples of the patient group in the form of the Hoehn & Yahr scale (N = 48) and disease duration (i.e., time since diagnosis; N = 56).

Neuroimaging data from 65 healthy older adults were collated from the PREVENT-AD (N = 50)^84^ and OMEGA (N = 15)^85^ data repositories to serve as a comparison group for the patients with PD. These participants were selected so that their demographic characteristics, including age (Mann-Whitney U test; *W* = 1551.50, *p* = .067), self-reported sex (chi-squared test; χ^2^ = 0.61, *p* = .434), handedness (chi-squared test; χ^2^ = 0.29, *p* = .863), and highest level of education (Mann-Whitney U test; *W* = 1831.50, *p* = .615), did not significantly differ from those of the patient group. Group demographic summary statistics and comparisons, as well as clinical summary statistics for the patient group, can be found in Table 1.

### Speech Sample Collection, Rating & Processing

Four auditory speech samples of cued sentence repetitions were recorded from each patient by the same neuropsychologist. Participants were fluent in French and/or English, and were allowed to hear and speak the sentences for repetition in the language of their choice (French: N = 51; English: N = 8). Speech samples were recorded an average of 19.63 days (SD = 53.44) from the date of neuroimaging data collection. All patients repeated four sentences (two easy, two hard) in the same order (see *Supplemenal Materials: Sentence Repetitions*). Sentences were taken from an in-house neuropsychological battery at the Montreal Neurological Institute, and were matched for difficulty and length. Sentences were pre-recorded by a female native speaker of each language and played to the participants at test time. Speech samples were recorded from participants using a Shure SM10ACN Cardioid Dynamic head-worn microphone. The microphone was positioned such that it was comfortable for the participants to wear, approximately at a two-finger distance to the participant’s mouth. Recordings were performed using a Tascam DR-10L Digital Audio Recorder.

The speech features of the recordings were then quantified in two ways: (1) using an automated extraction approach^4,86^, and (2) using the new Javascript toolbox *Audio-Tokens*^54^ to collect speech impairment ratings from non-experts. The automated extraction approach quantified the following features of the speech data using Praat^87^ with the *Python-parselmouth* interface^88^: harmonics-to-noise ratio (hnr), timing and amplitude fluctuations of glottal pulses (jitter and shimmer, respectively), standard deviation of pitch measured over voiced segments (f0_std), and a proxy for vowel space (area), namely the product of the inter-quartile range of F1 and F2 values measured over voiced segments (as a simplified version of the procedure described in Sandoval et al.^86^). To derive a single measure roughly representing voice quality from these features, the first principal component was extracted from the hnr, jitter, and shimmer features. The resulting metric, f0_std, and area were used in further analyses.

For the non-expert ratings of the speech samples, three individuals with little-to-no experience in speech analysis each rated multiple features of every sample on a continuous scale, including the magnitude of impairments in voice (instruction: “*Does the speaker’s voice sound harsh or breathy?*”), articulation (instruction: “*Is the speaker’s articulation slurred or imprecise?*”), and prosody (instruction: “*Does the speech sound flat or monotonous?*”). Sample ratings were performed in the *Audio-Tokens* toolbox^54^ (Fig. 1A), which allowed for interactive and dynamic comparison of speech samples across patients. The resulting values were then averaged across the 4 sentences for each speech feature/rater/patient, and the intraclass correlation coefficient (ICC; type [C,k]: multiple raters, two-way random effects, consistency)^89^ was computed to assess inter-rater reliability for each feature. Given the high consistency across raters for all three features (Fig. 1A), we used the mean of these values across the 3 raters to derive singular estimates of speech impairment for each feature in every patient.

### Magnetoencephalography Data Collection, Preprocessing & Analysis

Eyes-open resting-state MEG data were collected from each participant using a 275-channel whole-head CTF system (Port Coquitlam, British Columbia, Canada) at a sampling rate of 2400 Hz and with an antialiasing filter with a 600 Hz cut-off. Noise-cancellation was applied using CTF’s software-based built-in third-order spatial gradient noise filters. Recordings lasted a minimum of 5 min^90^ and were conducted with participants in the seated position as they fixated on a centrally-presented crosshair. Participants were monitored during data acquisition via real-time audio-video feeds from inside the shielded room, and continuous head position was recorded for each session.

MEG preprocessing was performed in *Brainstorm*^91^ unless otherwise specified, with default parameters and following good-practice guidelines^92^. The data were bandpass filtered between 1–200 Hz to reduce slow-wave drift and high-frequency noise, and notch filters were applied at the line-in frequency and harmonics (i.e., 60, 120 & 180 Hz). Signal space projectors (SSPs) were derived around cardiac and eye-blink events detected from ECG and EOG channels using the automated procedure available in *Brainstorm*^93^, reviewed and manually-corrected where necessary, and applied to the data. Additional SSPs were also used to attenuate highly-stereotyped artifacts on an individual basis. Artifact-reduced MEG data were then arbitrarily epoched into non-overlapping 6 second blocks and downsampled to 600 Hz. Data segments still containing major artifacts (e.g., SQUID jumps) were excluded within each session using the union of two standardized thresholds of ± 3 median absolute deviations from the median: one for signal amplitude and one for gradient. An average of 79.19 (SD = 14.68) epochs were used for further analysis (patients: 84.07 [SD = 7.78]; controls: 74.77 [SD = 17.82]). Empty-room recordings lasting at least 2 minutes were collected on or near the same day as the data recordings and were processed using the same pipeline, with the exception of the artifact SSPs, to model environmental noise statistics for source analysis.

MEG data were coregistered to each individual’s segmented T1-weighted MRI (Freesurfer *recon-all*)^94^ using approximately 100 digitized head points. For participants without useable MRI data (N = 11 patients with PD), a quasi-individualized anatomy was created and coregistered to the MEG data by warping the default Freesurfer anatomy to the head digitization points and anatomical landmarks for that participant^95^. Source imaging was performed per epoch using individually-fitted overlapping-spheres forward models (15,000 vertices, with current flows unconstrained to the cortical surface’s normal direction) and dynamic statistical parametric mapping (dSPM). Noise covariance estimated from the previously-mentioned empty-room recordings were included in the computation of the dSPM maps.

Inspired by previously-developed measures of multi-spectral neurophysiological signal pathology^96,97^ and cortical morphometric similarity in clinical populations^98,99^, we developed a new metric of neurophysiological spectral pathology derived from time-resolved MEG source maps: the Spectral Deviation Index (SDI; Fig. 2A). We computed vertex-wise estimates of power spectral density from the source-imaged MEG data using Welch’s method (3 s time window, 50% overlap), which we then averaged over canonical frequency bands (delta: 2–4 Hz; theta: 5–7 Hz; alpha: 8–12 Hz; beta: 15–29 Hz)^93^, and over all artifact-free 6-second epochs for each participant. The root-mean-square (RMS) norm of PSD across the three unconstrained orientations at each vertex location and for each participant was then projected onto a template cortical surface (*FSAverage*) for comparison across participants. For each PD participant, the resulting PSD map of spectrally-resolved estimates of neural power was correlated across frequencies (i.e., delta, theta, alpha and beta) at every spatial location (i.e., vertex) with the comparable estimates from each control participant. We generated for each patient the linearly-scaled SDI metric of spectral deviations per vertex by deriving the median of the resulting Pearson coefficients (*r*) across correlations with all control participants, normalizing these values using the Fisher transform (i.e., the inverse hyperbolic tangent; using the *atanh* function in Matlab), and subtracting them from 1 to generate a linearly-scaled metric of spectral deviation (i.e., higher values indicate greater functional neural pathology).

In addition to deriving individual SDI maps, we also used the source-imaged MEG data to investigate patterns of connectivity that relate to speech impairments in patients with PD. We extracted the first principal component from the three elementary source time series at each vertex location in each participant’s native space, and derived whole-cortex functional connectivity maps, using the peak vertex identified in our spatially-resolved SDI statistical analysis (back-transformed into each participant’s native space) as the seed. We used orthogonalized amplitude envelope correlations (AEC)^100,101^ as the connectivity measure, based on the same frequency definitions used for the SDI mapping. We estimated connectivity over each epoch and averaged the resulting AEC estimates across epochs, yielding a single AEC map per participant and frequency band. We projected these individual AEC maps onto the same template cortical surface (*FSAverage*) for group analyses.

Finally, we extracted several metrics to test for potential confounds of our primary effects of interest. To test whether our findings were mediated by local neurodegeneration, we estimated cortical thickness with the *recon-all* pipeline in *Freesurfer*^94^ and extracted values at the peak vertex of each significant statistical cluster for inclusion as nuisance covariates in post-hoc models. To determine whether SDI effects were related to shifts of the aperiodic broadband component of neural spectra, we processed PSDs with *specparam* (*Brainstorm* MATLAB version; frequency range = 2–40 Hz; Gaussian peak model; peak width limits = 0.5 –12 Hz; maximum n peaks = 3; minimum peak height = 3 dB; proximity threshold = 2 standard deviations of the largest peak; fixed aperiodic; no guess weight)^102^ to estimate the slope of aperiodic neural spectral components. We also investigated possible confound effects due to participant head motion, eye movements, and heart-rate variability: we extracted the RMS of signals from the head position indicators, EOG, and ECG channels, respectively. Alongside age, these derivations were included in *post hoc* statistical models to examine the robustness of the initial effect(s) of interest against these potential confounds.

### Statistical Analyses

We assessed relationships between continuous variables using the *lm* function in *R*^103^, with a significance threshold of *p* < .05. Model comparisons were performed using the Akaike information criterion (AIC), with differences in AIC (ΔAIC) between tested models of ΔAIC > 2 considered as meaningful^104^. Where appropriate^105^, we performed mediation analyses using a non-parametric bootstrapping approach for indirect effects with 10,000 simulations^106^. All statistical models included age as a nuisance covariate. Participants with missing data were excluded pairwise per model.

We performed statistical comparisons using spatially-resolved neural data, covarying out the effect of age, using *SPM12*. Initial tests used parametric general linear models to investigate relationships with speech impairment ratings (i.e., multiple regression with voice, prosody, and articulation impairment ratings as predictors), beyond the effects of age. Contrasts for each speech feature were thus corrected for age and independent of the other features. We used Threshold-Free Cluster Enhancement (TFCE; E = 1.0, H = 2.0; 5,000 permutations)^107^ to correct the resulting *F*-contrasts for multiple comparisons across vertices. TFCE avoids the assumptions of parametric modeling, accounts for potential non-uniform spatial autocorrelation of the data, and avoids the arbitrary selection of cluster-defining thresholds. We applied a final cluster-wise threshold of *p*FWE < .05 to determine statistical significance, and used the TFCE clusters at this threshold to mask the original statistical values (i.e., vertex-wise *F* values) for visualization. We extracted data from the vertex exhibiting the strongest statistical relationship in each cluster (i.e., the “peak vertex”) for subsequent analysis and visualization.

To determine the relative contribution of individual frequency bands to the significant SDI-speech relationships, we used an adapted leave-one-out approach. We recomputed SDIs at each peak-vertex four times – each time excluding data from one frequency band. Speech impairment ratings were then regressed on these modified SDI values, and we derived the ΔAIC between each leave-one-out model and the original model, with higher values indicating greater contribution to the original effect. Lines-of-best-fit were also fitted to the data *post hoc* and plotted to display the nature of the underlying relationships between spectral power and speech ratings for each frequency band.

## Data Availability

All data produced in the present study are available upon reasonable request to the authors

## Acknowledgements

This work was supported by grant F32-NS119375 to AIW from the United States National Institutes of Health (NIH); to PD from the Richard & Edith Strauss Foundation; to EAF as a Foundation Grant from the Canadian Institutes of Health Research (CIHR; FDN-154301) and the CIHR Canada Research Chair (Tier 1) of Parkinson’s Disease; to DK from the Healthy Brains for Healthy Lives (HBHL) initiative, the Natural Sciences and Engineering Research Council of Canada, the Centre for Research on Brain, Language and Music, the Edith Strauss Foundation, and a private donor; and to SB from by a NSERC Discovery grant, the Healthy Brains for Healthy Lives initiative of McGill University under the Canada First Research Excellence Fund, the CIHR Canada Research Chair (Tier 1) of Neural Dynamics of Brain Systems and the NIH (1R01EB026299). Data collection and sharing for this project was provided by the Quebec Parkinson Network (QPN), the Pre-symptomatic Evaluation of Novel or Experimental Treatments for Alzheimer’s Disease (PREVENT-AD; release 6.0) program, and the Open MEG Archives (OMEGA). The funders had no role in study design, data collection and analysis, decision to publish, or preparation of the manuscript.

The QPN is funded by a grant from Fonds de recherche du Québec - Santé (FRQS). PREVENT-AD was launched in 2011 as a $13.5 million, 7-year public-private partnership using funds provided by McGill University, the FRQS, an unrestricted research grant from Pfizer Canada, the Levesque Foundation, the Douglas Hospital Research Centre and Foundation, the Government of Canada, and the Canada Fund for Innovation. Private sector contributions are facilitated by the Development Office of the McGill University Faculty of Medicine and by the Douglas Hospital Research Centre Foundation (http://www.douglas.qc.ca/). OMEGA and the Brainstorm app are supported by funding to SB from the NIH (R01-EB026299), a Discovery grant from the Natural Science and Engineering Research Council of Canada (436355-13), the CIHR Canada research Chair in Neural Dynamics of Brain Systems, the Brain Canada Foundation with support from Health Canada, and the Innovative Ideas program from the Canada First Research Excellence Fund, awarded to McGill University for the HBHL initiative.

