## Supplementary material for "Aberrant neurophysiological signaling underlies speech impairments in Parkinson’s disease": Supplemenal Materials

### Supplemental Materials & Methods

#### *Sentences for Repetition*

English:
- *The mother washes her child*. (easy)
- *Her hair got tangled in the wind.* (easy)
- *I can't remember where I put my brand new bag* (hard)
- *They caught some trout while they were fishing on the lake* (hard)

French:
- *Je cherche un nouveau manteau.* (easy)
- *Je veux le plus petit morceau.* (easy)
- *Les chevaux ont eu très peur lors de l'incendie.* (hard)
- *Le gros chien a attaqué l'homme très soudainement.* (hard)

### Supplemental Figures

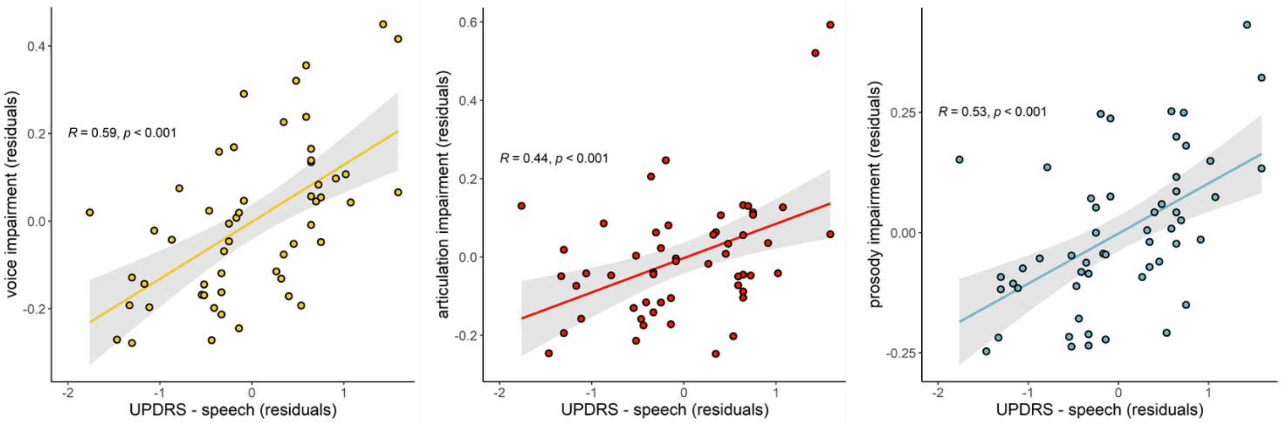


**Figure S1. Non-expert speech impairment ratings are predicted by clinical motor speech assessments.** Significant linear relationships between voice, articulation, and prosody impairments and the speech sub-score of the Unified Parkinson's Disease Rating Scale part III. All models controlled for age. Shaded intervals represent the 95% confidence interval.
